# Characteristics of Highly Creative Surgeons (The INSPIRE Study): An International Mixed-Methods Study Protocol

**DOI:** 10.64898/2026.05.15.26353308

**Authors:** Alex Thabane, Tyler McKechnie, Phillip Staibano, Cristian Scheau, Serban Dragosloveanu, Ernesto Guerra Farfan, Roger Rojas Sayol, Vikram Arora, Goran Calic, Sameer Parpia, Jason W. Busse, Nouf Hamoudi, Diya Patel, Roni-Reiter Palmon, Mohit Bhandari

## Abstract

**Introduction:** Creativity is important in surgery for problem-solving in the operating room and the development of surgical innovations that improve patient outcomes. However, our limited understanding of what the characteristics and competencies of the highly creative surgeon are has inhibited our ability to develop the tools, programs and interventions necessary for cultivating the creativity of surgeons. We present the protocol for the INSPIRE Study, which aims to identify the factors associated with high creative achievement in surgeons.

**Methods and Analysis:** We have designed a sequential mixed-method study, including a cohort study accompanied by qualitative semi-structured interviews. The primary objective of this study will be to identify factors associated with high creative achievement in surgeons, to be assessed through direct involvement in innovation or invention, or a top score (10 out of 10) on any domain in the Inventory of Creative Activities and Achievements questionnaire. We plan to measure 39 different personal, domain-specific, domain-general, and environmental/motivational variables, chosen based on previous literature and on exploratory grounds, to be assessed as possible factors of creative potential. Multivariable logistic regression is planned, with high creative achievement as the dependent variable and all 39 potential factors of creative potential as independent variables.

**Ethics and Dissemination:** Ethics approval from the Hamilton Integrated Research Ethics Board has been obtained and no harm is expected due to participation in this study. To facilitate knowledge translation, we plan to publish the feasibility data and results in peer-reviewed journals, and present at international surgical and creativity conferences.

## INTRODUCTION

The world is changing. As our daily life and work evolves in response, identifying and cultivating the skills required to survive and thrive within is an important societal task. Creativity, the ability to generate novel and useful ideas (1, 2), has consistently been identified as a critical 21^st^ century skill (3-5).

The ability to be creative is colloquially associated with artists and musicians, but its value and manifestation extends beyond the traditional arts: creativity can be used to solve problems, create new products and services, and drive economic growth (6). In recent years there has been a growing evidence-base supporting the need for creativity across a range of domains (7-9) – including surgery (10).

On the surface, surgery is a profession highly unamenable to creativity: it is highly risk-averse and subsequently constrained through guidelines, best practices, and regimented instruction. But throughout its history, surgeons, and the patients they treat, have profited from creative ideas: surgical anesthesia, percutaneous endoscopic gastrostomy, and minimally invasive surgery are just a few novel and useful ideas that have revolutionized patient care for the better (11-15). Further, creativity is important for problems for which there are no well-defined solutions (i.e., ill-defined problems), which makes it a particularly useful ability when unfamiliar, emergency situations arise in the operating room (16, 17).

Given its value and importance in surgery, and with very little scientific literature on the topic (18), there have been recent efforts to explore the nature and distribution of creativity in surgery. This includes the generation of a surgery-specific definition of creativity, providing the necessary theoretical foundation for valid and reliable scientific study (10), and a mixed-methods survey of divergent thinking which found evidence of below-average idea originality among surgeons and a negative association between divergent thinking and surgical training (19). These preliminary studies indicate a sort of “creativity crisis” in surgery – and the possible need for new approaches to surgical training that encourage creative capacities instead of impeding them.

Precluding the development of valid and effective tools, interventions, and programs improving the creativity of surgeons is our limited understanding of what exactly makes a surgeon creative. Most studies of creativity, including the previously mentioned mixed-method study, measure creativity with tests of divergent thinking (20, 21). However, divergent thinking alone cannot be considered a holistic measure of creative potential – personality, general cognitive ability, motivation, and domain-specific competencies are factors that likely contribute to a surgeon’s creative potential (22-27). More, the existing literature on creativity in surgery has exclusively been conducted in Western countries, which limits the generalizability of the available evidence and prevents any exploration of differences in creativity across regions and cultures; current evidence indicates that that perceptions of creativity, and performance on creativity assessments, differ by culture (28, 29). There is a clear need for a large-scale and global study of creativity in surgeons, involving a robust and multi-dimensional approach to creativity assessment, to fully characterize highly creative surgeons, including their background, characteristics, and competencies.

With such a study in mind, we present a protocol for the INternational, proSPective cohort study of creativIty in suRgEons (INSPIRE Study), a longitudinal, mixed-methods study of creativity in surgery which upon completion will represent the largest international study of creativity among surgeons.

## METHODOLOGY

This study has received research ethics approval from the Hamilton Integrated Research Ethics Board (#18519) and has been registered on Open Science Framework (30).

### Study Objective(s)

The primary objective of this study will be to identify the personal, domain-specific, domain-general, and environmental factors associated with high creative achievement in surgeons.

As secondary objectives, we plan to:

A. Assess the feasibility of the quantitative survey;
B. Explore whether divergent thinking and creative self-efficacy change over time; and
C. Explore surgeons’ perceptions of creativity, including
  i. How it is conceptualized and defined,
  ii. Its importance for surgical problem-solving and innovation,
  iii. The effect of surgical training on creativity, and
  iv. Differences in perceptions of creativity by cultural context

### Study Design

This study will employ a sequential, explanatory mixed-methods design, to be conducted in three stages. We have chosen a mixed-method design on grounds of complementarity, with the results from the qualitative portion being used to clarify and explain the quantitative findings.

The initial quantitative phase includes a 5-year longitudinal survey of creativity, with an initial feasibility assessment in the first stage, followed by the full launch of the survey in a second stage. The subsequent qualitative phase represents the third stage of the study and will involve qualitative interviews with surgeons identified during the quantitative phase to have accomplished high creative achievements. We will use pragmatism as the philosophical stance for mixing quantitative and qualitative methodologies (31), with a focus on combining methods of data collection for ‘what works best’ in answering the research question (32).

### Steering Committee

This study will be led by an international and interdisciplinary steering committee of seven members with extensive experience designing and conducting large, international studies in the field of psychology and medicine. This includes a PhD candidate in health research methodology leading a research program on creativity (AT), a Professor of Strategic Management with a background in creativity and cognitive neuroscience (GC), a Professor of Anesthesia with extensive expertise in research methods (JWB), a Professor of Biostatistics with experience designing large international studies (SP), a Distinguished Professor of Industrial/Organizational Psychology with over 30 years conducting creativity research (RRP), two surgeon-residents with PhD-level training in research methods (TM; PS), and one Distinguished Professor and national research chair in surgical innovation with over 30 years of experience conducting large, international studies in surgery (MB). The committee will be co-chaired by AT and MB.

### Theoretical Assumptions

For the purposes of this study, we conceptualize creativity in surgery as the generation of novel and useful products, ideas, and solutions that leads to improved patient outcomes (10). In alignment with current theories of creativity (10, 22, 25-27), we conceptualize creative potential to be a multicomponent construct involving a variety of characteristics and competencies such as intelligence, knowledge, domain-relevant skills, personality, and motivation.

### Quantitative Sampling Technique & Recruitment

For the quantitative phase of this study, we will conduct non-probabilistic convenience sampling of any surgeon who has completed or is currently completing an accredited surgical training program in their respective country. We will exclude surgeons who self-report as unable to read, write, or understand English, or refuse to consent for any other reason.

To facilitate the recruitment of a large, heterogeneous, and international sample of surgeons, we have planned a multi-modal recruitment strategy. First, we plan to send personalized emails to surgeons in the personal networks of our steering committees inviting them to participate in the study. Second, we plan to invite local, national, and international surgical associations, surgical departments, and surgical divisions with email lists to circulate the survey their constituents. Third, we plan to post information and links to the survey on the LinkedIn and X platforms. Lastly, we also plan to recruit participants through word of mouth at conferences, clinical rounds, and other settings where surgeons may be present.

### High Creative Achievement

To assess high creative achievement, defined as the real-world achievement of highly novel and useful products, ideas, or solutions, we will consider lead involvement in innovation or invention, or a top score on a validated measure of creative achievement, to be indicative of high creative achievement.

First, we plan to ask the following binary response questions: “Have you ever directly ideated and developed an innovation, including but not limited to a new product, procedure, device, or process?”; and “Have you ever received a patent for an invention?”. Both questions will be followed up with open-text fields to describe the details of the innovation or invention. A surgeon will be considered to have accomplished a high creative achievement in the case of affirmative response to either question. Second, we will use an adapted version of the Inventory of Creative Activities and Achievements (ICAA) questionnaire (33) to assess creative achievement across eight domains (Literature, Music, Arts & Crafts, Cooking, Sports, Visual Arts, Performing Arts, and Science & Engineering). Participants will self-rate their highest attained level of creative achievement for each domain, rated on a scale from 0 (I have never engaged in any of these domains) to 10 (I have sold some of my work in these domains). Participants with a high level of creative achievement in any domain (a score of 10 out of 10) will also be considered to have accomplished a high creative achievement.

### Potential Factors of Creative Potential

To identify factors associated with high creative achievement (i.e., factors of creative potential), we plan to collect the following the personal, domain-specific, domain-general, and environmental factors, chosen based on the current literature (22-24, 34-43) and on exploratory grounds:

#### Personal Factors

We will evaluate 16 personal factors. The following personal factors will be self-reported by the surgeon – age; biological sex; country of birth; country of surgical practice; ethnicity; first spoken language; bilingual status; immigration status (i.e., 1^st^ generation, 2^nd^ generation, 3^rd^+ generation), family history of invention, genius, or creative achievement; presence of creative friends; presence of creative colleagues; creative hobbies; medications; travel frequency; number of countries visited; and whether the surgeon makes time to think about or concentrate on new ideas or solutions to problems (i.e., “think time”) (44).

#### Professional Factors

We will evaluate 15 professional factors. The following professional factors will be self-reported by the surgeon – surgical specialty; years of surgical experience (post-medical degree); number of grants; total amount of grant money received; number of publications; H-index; highest academic degree (excluding medical degree); yearly salary; career stage (i.e., early career [0-5 years], mid-career [5-15 years], late career [15+ years]); number of patents; number of companies started; highest held leadership role (e.g., dean, department chair, head of surgical division); whether the surgeon has a creative mentor, and; setting of surgical practice (hospital vs. clinic; academic vs private).

#### Environmental Factors

We will evaluate 3 environmental factors. The following statements on environmental characteristics will be rated on a 5-point Likert scale (1 = strongly disagree to 5 = strongly agree) – “creativity was encouraged in my family”; “creativity is encouraged in my country”; and “creativity is encouraged by my institution and its leadership”.

#### Motivation

To assess creative motivation to engage in creative behaviours, we used the nine-item Creativity Motivation Scale (CMS), which has demonstrated acceptable reliability in several countries (α=0.77-0.88) (45).

#### Openness to Experience

We plan use a 10-item scale to measure openness to experience from the International Personality Item Pool (IPIP) (46), derived from the Revised NEO Personality Inventory (47). Scores will range from 10 (low openness) to 50 (high openness). The scale has demonstrated acceptable reliability (α=0.82).

#### Divergent Thinking

To measure divergent thinking, we plan to administer the Alternate Uses Task (AUT) (48), one of the most popular measures of creativity potential and creative cognition. The AUT presents the participant with a well-known object and asks them to list as many “alternate uses”. Following best practices and previous research (49, 50), we will give participants 3 minutes to generate responses, and instruct participants to “be creative” and describe the task with the following prompt:

> *For this task, you’ll be asked to come up with as many original and creative uses for a* ***shoe*** *as you can. The goal is to come up with creative ideas, which are ideas that strike people as clever, unusual, interesting, uncommon, humorous, innovative, or different*.
>
> *Your ideas don’t have to be practical or realistic; they can be silly or strange, even, so long as they are CREATIVE uses rather than ordinary uses*.
>
> *You can enter as many ideas as you like. The task will take 3 minutes. You can type in as many ideas as you like until then, but creative quality is more important than quantity. It’s better to have a few really good ideas than a lot of uncreative ones. List as many ORIGINAL and CREATIVE uses for a shoe*.

Scoring of the AUT will be performed using the Ocsai 1.6 model, which is a multi-lingual, multi-task LLM model designed for assessing AUT items (51). This will include a fluency score (number of responses) and originality score (novelty of responses).

#### Creative Self-Efficacy

To measure creative self-efficacy, defined as “the belief that one has the ability to produce creative outcomes” (52), we will use the using the Short Scale of Creative Self (SSCS) (53). The SSCS assesses creative self-efficacy through 11 statements assessed with a 5-point Likert scale (‘definitely not’ to ‘definitely yes’) which the participant scores based on how well it describes them. The SSCS has been found to have strong psychometric properties, including uni-dimensionality, reliability, and item discrimination (54).

We provide a complete list of the independent variables, and hypotheses of association with high creative achievement, in **S1 Appendix**.

### Survey Administration & Follow-Up

Before initiating the first stage of the survey, we will pre-test the survey with five surgeons to estimate how long the survey will take, as well as assess its face validity, comprehensiveness of content, interpretability (each to be rated on a 7-point Likert scale).

The final survey will be sent to participants electronically using an online survey platform and consist of 3 pages. The first page will provide a short description of the study, a “commitment check”, asking participants whether they commit to providing thoughtful and accurate survey responses, and a collection of demographic information. The second page will involve the assessment of high creative achievement and the potential factors of creative potential. The third page will contain the AUT, programmed to automatically submit the participant responses after 3 minutes have passed.

Following completion of the initial survey, participants will be moved to a follow-up list. At 1-, 3- and 5-years follow-up, we will re-assess high creative achievement, the professional factors, divergent thinking, and creative self-efficacy.

### Feasibility Outcomes

For the first 4 months of the study, we plan to assess the feasibility of the quantitative survey to determine whether modifications will be required before initiating the second stage. We plan to estimate the following outcomes:

1. Response rate, expressed as the proportion of surgeons who complete at least one page of the survey among those who have received an invitation to participate.
2. Recruitment rate, expressed as the number of surgeons who completed at least one page of the survey per month. We will calculate an overall recruitment rate, as well as the rate of recruitment by each recruitment approach (i.e., personalized email, group list, word of mouth, social media).
3. Completion rate, expressed as the proportion of surgeons who fully complete the survey among those who complete the first page of the survey.
4. “High creative achievement” rate, expressed as the proportion of surgeons who meet the criterion for “high creative achievement” among those who fully completed the survey.

We have set “traffic light” criteria for each feasibility outcome (55), an approach successfully used in previous feasibility studies (56, 57): a “green light” indicates that the survey can move forward without any modifications due to this outcome, a “yellow light” indicates that some modifications are necessary, and a “red light” that serious modifications are required. The criteria are outlined in **Table 1**.

**Table 1:**
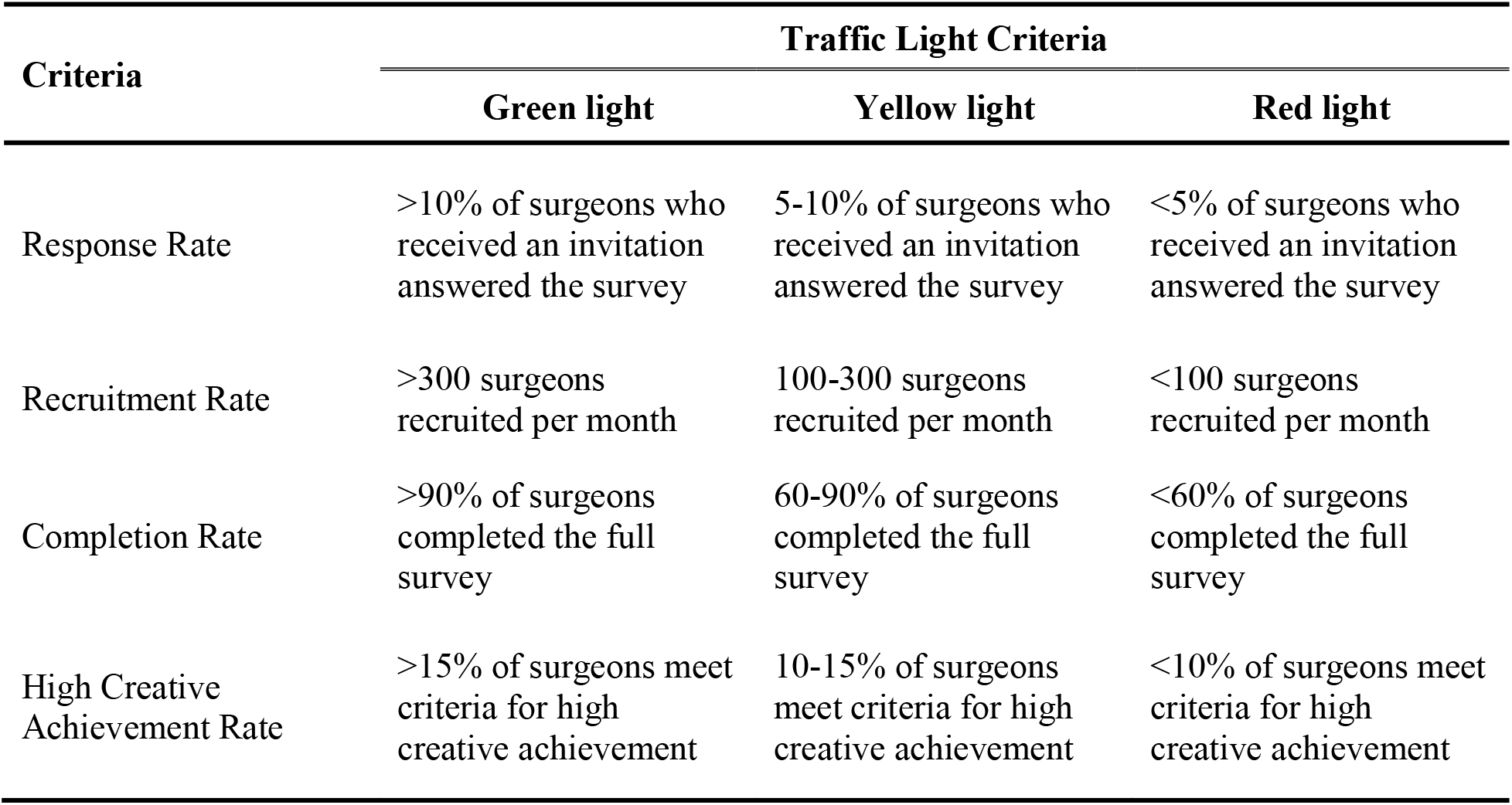
Traffic Light Criteria for Feasibility Outcomes.

| Criteria | Traffic Light Criteria |  |  |
| --- | --- | --- | --- |
|  | Green light | Yellow light | Red light |
| Response Rate | >10% of surgeons who received an invitation answered the survey | 5-10% of surgeons who received an invitation answered the survey | <5% of surgeons who received an invitation answered the survey |
| Recruitment Rate | >300 surgeons recruited per month | 100-300 surgeons recruited per month | <100 surgeons recruited per month |
| Completion Rate | >90% of surgeons completed the full survey | 60-90% of surgeons completed the full survey | <60% of surgeons completed the full survey |
| High Creative Achievement Rate | >15% of surgeons meet criteria for high creative achievement | 10-15% of surgeons meet criteria for high creative achievement | <10% of surgeons meet criteria for high creative achievement |

### Quantitative Analyses

We have planned descriptive statistics to describe the demographic characteristics of the sample, reported as means (with standard deviations) for continuous outcomes when normally distributed, or as medians (with inter-quartile ranges) when not, and counts (with percentages) for categorical outcomes.

To meet our primary objective of identifying factors associated with highly creative surgeons, we have planned multivariable logistic regression with a dependent variable of “high creative achievement”, and all 39 factors of creative potential as independent variables. We will perform an assessment of model assumptions and goodness-of-fit by examining the residuals. We will also explore variance inflation factors (VIFs) to assess collinearity between independent variables. If multicollinearity is detected between two or more variables (i.e., VIFs ≥5) (58), then we will construct multiple regression models, each containing one of the collinear variables, comparing them to one another and selecting the model which produces the lowest Akaike information criterion. The results of the regression analyses will be reported as odds ratios (ORs), with corresponding 95% confidence intervals (CIs) and the associated p-value. We plan to perform all analyses with R Software (version 4.3.2) via RStudio.

For secondary objective of assessing study feasibility, we will calculate the response rate, completion rate, and high creative achievement rate as counts (with percentages). The recruitment rate will be reported as a rate (i.e., number of recruited surgeons per month). These results will be evaluated against the traffic light criteria in **Table 1** and reported in a colour coded table.

For secondary objective of exploring the change divergent thinking and creative self-efficacy over time, we also plan three separate repeated measures ANOVA analyses for SSCS scores, AUT fluency and AUT originality scores across all time-points (baseline, 1-year, 3-years, 5-years), to be conducted upon completion of the study. We will perform assessments of model assumptions and report the results of the statistical testing as F-values with associated p-values. In the case of a statistically significant ANOVA test, we will perform *post-hoc*, Bonferroni corrected t-tests to identify which timepoints differed among one another.

Only participants who passed the commitment check will be included in the analyses.

### Quantitative Sample Size Calculation

To adequately power our study for the primary objective, we estimated the proportion of surgeons who will have accomplished a “high creative achievement”: ICAA results are typically highly positively skewed (43), which is indicative of a low rate of creative achievement from both a domain-specific and domain-general perspective. Assuming that 15% of participants will meet the requirements for “high creative achievement”, and using an events per variable ratio of ten for the 39 potential factors of creative potential included as independent variables in the regression model (59), we estimate a necessary sample size of 2,600 surgeons (to recruit 390 high creative achievement surgeons at a rate of 15%).

### Qualitative Sampling Technique & Recruitment

We plan to invite surgeons that met the criteria for high creative achievement during the quantitative phase for semi-structured interviews via email. We plan to use a purposive maximum variation sampling approach to ensure a range of perspectives and sociodemographic characteristics (60).

### Semi-Structured Interviews

We will conduct semi-structured interviews with all consenting surgeons one-on-one in a conversational manner, asking both closed and open-ended questions to facilitate a guided yet exploratory discussion on creativity (61). The following interview guide questions will be used:

1. How would you define or conceptualize creativity?
2. Do you think creativity is important in surgery?
  a. In what ways (for example, in the operating room or for surgical innovation)?
3. What effect, if any, do you think surgical training has had on your creativity?
4. Do you think the survey you completed accurately assesses creative ability or achievement? Why or why not?
5. What do you think an intervention for enhancing surgeon creativity should consider or include?

The interview will be conducted in person or over the Zoom platform depending on the availability of the interviewee.

Field notes will be taken during the interviews to document emergent themes and other observations. Trustworthiness of our qualitative data will be assessed via member checking (i.e., participants will be sent raw transcripts of their interviews as well as a summary of the major themes to be reviewed and affirmed) (32). In addition, a reflexive journal and audit trail will be maintained to track potential researcher bias and document decisions made around qualitative data collection and analysis procedures.

### Qualitative Analyses

We plan to perform a thematic analysis using a post-positivist approach, which aims to achieve objectivity and reliability in the analysis, minimizing researcher bias (62). A team of two researchers with training in qualitative research methods will conduct the coding and analyses using the relevant software (i.e., NVivo). The themes and patterns that emerge from the data will be organized and narratively described.

### Qualitative Sample Size Calculation

Utilizing sample size rules of thumb sourced from the qualitative research literature, we estimate that we will require a minimum of 12 participants to reach the point of data saturation (63). Data saturation will be calculated as the point at which no new themes in the data set emerge for 3 interviews in a row.

### Study Schedule / Timeline

Recruitment is scheduled to begin in November of 2025. We expect the feasibility phase to be complete by April of 2026, and to reach our full sample size of 2,600 surgeons by December of 2026. The quantitative interview phase will likely take 1 year to complete and will happen concurrently with cohort follow-up. All objectives are estimated to be complete by December 2031.

## ETHICS & DISSEMINATION

### Ethical Considerations

This proposed study has received ethics approval from the Hamilton Integrated Research Ethics Board (Project #18519). There is no expected risk in the participation of a study of this nature, however, patients may feel stressed or anxious given the timed nature of the AUT. An implied consent model will be utilized for the survey portion of the study, and written consent will be sought for potential interview participants prior to conducting the interview.

### Knowledge Translation

We aim to disseminate the results and implications of our study in several ways. In addition to the publication of this protocol, we aim to publish the results of the feasibility data by mid-2026, the results of the primary objective analyses by early 2027, and the results of remaining secondary objective analyses by the end of the study period. The results of these papers will be submitted to surgical and creativity conferences for presentation, and efforts will be made to communicate the results locally, nationally, and internationally.

## CONCLUSION

The ability to be creative in surgery can improve patient outcomes through enhanced problem-solving ability and the generation of new surgical innovations. This mixed-methods study will provide a multidimensional and international audit of creativity among surgeons, to be used to develop future tools, interventions, and programs to cultivate the creativity of surgeons.

## Data Availability

As a protocol, this paper does not contain any participant data.

