## Supplemental Appendix 1 for "Characteristics of Highly Creative Surgeons (The INSPIRE Study): An International Mixed-Methods Study Protocol"

| **Variable** | **Hypothesized Relationship with High Creative Achievement** |
| --- | --- |
| **Personal** **Factors** | |
| Age | No Association |
| Biological Sex | No Association |
| Country of Birth | No Association |
| Country of Surgical Practice | No Association |
| Ethnicity | No Association |
| First Spoken Language | No Association |
| Bilingual Status | Positive Association |
| Immigration Status | Positive Association |
| Family History of Creative Achievement | Positive Association |
| Presence of Creative Friends | Positive Association |
| Presence of Creative Colleagues | Positive Association |
| Creative Hobbies | Positive Association |
| Medication | No Association |
| Travel Frequency | Positive Association |
| Number of Countries Visited | Positive Association |
| “Think Time” | Positive Association |
| **Professional Factors** | |
| Surgical Specialty | No Association |
| Years of Surgical Experience | Negative Association |
| Number of Grants | No Association |
| Total Grant Money Earned | No Association |
| Number of Publications | No Association |
| H-index | No Association |
| Highest Academic Degree | No Association |
| Yearly Salary | No Association |
| Career Stage | No Association |
| Number of Patents | Positive Association |
| Number of Companies Started | Positive Association |
| Highest Held Leadership Role | No Association |
| Creative Mentor | Positive Association |
| Surgical Setting (Clinic vs. Hospital) | No Association |
| Surgical Setting (Academic vs. Private) | Private – Positive Association |
| **Motivational/Environmental Factors** | |
| Creativity Encouraged in Family | Positive Association |
| Creativity Encouraged in Country | Positive Association |
| Creativity Encouraged by Institution & Leadership | Positive Association |
| Creative Motivation | Positive Association |
| **Personality** | |
| Openness to Experience | Positive Association |
| **Intelligence** | |
| Divergent Thinking – Fluency | No Association |
| Divergent Thinking – Originality | Positive Association |
| **Self-Perception** | |
| Creative Self-Efficacy | Positive Association |
